# Climate change impacts on Zika and dengue risk in four Brazilian cities: projections using a temperature-dependent basic reproduction number

**DOI:** 10.1101/2022.09.26.22280352

**Authors:** Hannah Van Wyk, Joseph NS Eisenberg, Andrew F. Brouwer

## Abstract

For vectorborne diseases the basic reproduction number *ℛ*_0_, a measure of a disease’s epidemic potential, is highly temperature dependent. Recent work characterizing these temperature dependencies has highlighted how climate change may impact geographic disease spread. We extend this prior work by examining how newly emerging diseases, like Zika will be impacted by specific future climate change scenarios in four diverse regions of Brazil, a country that has been profoundly impacted by Zika. We estimated a temperature-dependent *ℛ*_0_(*T*), derived from a compartmental transmission model, characterizing Zika (and, for comparison, dengue) transmission potential. We obtained historical temperature data for the 5-year period 2015–2019 and projections for 2045–2049 by fitting cubic spline interpolations to data from simulated atmospheric data provided by the CMIP-6 project (specifically, generated by the GFDL-ESM4 model), which provides projections under four Shared Socioeconomic Pathways (SSP). These four SSP scenarios correspond to varying levels of climate change severity. We applied this approach to four Brazilian cities (Manaus, Recife, Rio de Janeiro, and São Paulo) that represent diverse climatic regions. Our model predicts that the *ℛ*_0_(*T*) for Zika peaks at 2.7 around 30°C, while for dengue it peaks at 6.8 around 31°C. We find that the epidemic potential of Zika and dengue will increase beyond current levels in Brazil in all of the climate scenarios. For Manaus, we predict that the annual *ℛ*_0_ range will increase from 2.1–2.5, to 2.3–2.7, for Recife we project an increase from 0.4–1.9 to 0.6–2.3, for Rio de Janeiro from 0–1.9 to 0–2.3, and for São Paulo from 0–0.3 to 0–0.7. As Zika immunity wanes and temperatures increase, there will be increasing epidemic potential and longer transmission seasons, especially in regions where transmission is currently marginal. Surveillance systems should be implemented and sustained for early detection.

**Author summary:** Rising temperatures through climate change are expected to increase arboviral disease pressure, so understanding the impact of climate change on newly emerging diseases such as Zika is essential to prepare for future outbreaks. However, because disease transmission may be less effective at very high temperatures, it is uncertain whether risk will uniformly increase in different regions. Mathematical modeling is a useful tool for predicting the impact of temperature on arbovirus risk. We used a temperature-dependent infectious disease transmission model to derive a temperature-dependent basic reproduction number. We then used historical temperature data and temperature projections for the years 2045-2049 to forecast Zika risk in four cities in Brazil under various climate change scenarios. We predict an overall increase in arbovirus risk, as well as extended risk seasons in cities that are not currently suitable for year-round spread, such as Rio de Janeiro. We also found little-to-no protective effect of increasing temperatures even in warmer climates like Manaus. Our results indicate that preparation for future Zika outbreaks (and of those of other arboviruses including dengue) should include the implementation of national disease surveillance and early detection systems.

## Introduction

The Zika and dengue viruses are closely related arboviruses that are primarily transmitted to humans through the *Aedes aegypti* and *A. albopictus* mosquitoes. Brazil carries an especially large share of the disease burden, with an estimated 1.5 million Zika cases since the beginning of the 2015–16 outbreak [1]. Zika was introduced in the Americas in 2015 [2], causing numerous outbreaks in countries throughout Latin America, including Brazil, Colombia, and Venezuela. Because vector-borne disease transmission depends on temperature, recent work has outlined the potential for climate change to facilitate its re-emergence (and emergence in new regions) [3–5]. Given the concerning health outcomes of Zika —including microcephaly and Guillain-Barre syndrome—the unpredictability of how the changing climate will influence the spread of the virus throughout the western hemisphere is a growing cause of concern.

Dengue has a longer history in the region, originally emerging in the Americas in the 1600s [6]. It was eliminated by the 1960s through widespread use of pesticides, but it re-emerged in the early 1980s [7]. Since its re-emergence, dengue has remained endemic throughout many Latin American countries [8]. Due to dengue’s endemicity and wide geographic spread, it has been better studied than Zika and provides a useful point of comparison as we consider the potential impact of climate change on these arboviruses.

As a result of climate change, it is estimated that about half of the world population will live in geographic regions that will be suitable for arbovirus transmission by the year 2050 [9]. Several factors make Brazil particularly vulnerable to both the drivers and impacts of climate change. Primary among these is the deforestation within the Amazonian region as well as widespread increases in temperature, both of which are conducive to mosquito breeding [10]. In fact, the 2015 Zika outbreak in Brazil has been partially attributed to the El Niño conditions that year [11]; *Aedes aegypti*, the primary vector of Zika and dengue [12], is particularly suited to warm, humid conditions. Brazil, therefore, is an important region to study forecasts of Zika transmission potential. Prior forecast studies of arbovirus transmission potential were primarily focused on North America [13–15] with fewer studies occurring in South America.

In the 1990s, when researchers started using mathematical modeling to consider the impacts of climate change on vector-borne disease transmission, several studies began to incorporate temperature-dependent parameters such as vector competence, vector lifespan, and extrinsic incubation period [5, 16]. More recently, temperature-dependent *ℛ*_0_s for vector-borne diseases have revealed an interesting range of peaks depending on the pathogen and mosquito species [17–19]. Of particular interest, Mordecai et al. found that disease risk peaks at the highest temperatures for pathogens that are transmitted by the *A. aegypti* mosquito [19]. The same group has also theorized that this finding means that with increasing temperatures, vector-borne disease risk in Africa will shift from malaria to arboviral diseases [20]. As climate change has the potential to shift much of the world into temperatures where these higher peaks occur, it is important to better understand both the range of uncertainty across climate change scenarios as well as the likely geographic and temporal heterogeneity in disease risk.

We extend prior work [21] that developed temperature dependent *ℛ*_0_ expressions to forecast future global trends of Zika and, as a comparison, dengue transmission risk in Brazil for the years 2045–2049, across a range of plausible climate change scenarios. Specifically, we explore how projections might vary across regions within a country and the likely impact of year-to-year temperature variation. We developed a basic reproduction number *ℛ*_0_(*T*) as function of temperature-dependent vector parameters, and we used this temperature-dependent *ℛ*_0_(*T*) to project seasonal disease risk in four Brazilian cities representative of the different climate regions of Brazil. Our work extends and complements existing projections of Zika risk in Brazil [4] and the temperature-dependent reproduction number literature more broadly [17–21] by assessing geographic and year-to-year heterogeneity in projected risk across climate change scenarios.

## Materials and methods

### Data

To examine the potential impacts of climate change across a variety of climates, we selected four cities representative of diverse climatic regions of Brazil: Manaus, a city in the Amazon Rainforest with a tropical rainforest climate; Recife, an Atlantic coastal city with a tropical monsoon climate; Rio de Janeiro, an Atlantic coastal city with a tropical savanna climate; and São Paulo, a southern city with a humid subtropical climate. All cities are at approximately sea level and within the suitable elevation range for an abundant *A. aegypti* population, i.e., up to 1,600 meters [22, 23].

We obtained the historical and projected future temperature (using the tas variable, or daily-mean 2-meter air temperature) data from ISIMIP (The Inter-Sectoral Impact Model Intercomparison Project) [24] from the downscaled bias-adjusted GFDL-ESM4 (Geophysical Fluid Dynamics Laboratory, NOAA) climate forcing [25–27]. GFDL-ESM4 is a CMIP-6 model (Coupled Model Intercomparison Project, phase 6), generally agreed to accurately capture historical temperatures in South America [28, 29]. To extract temperature data for each of the four cities, we calculated the nearest model grid point to each city’s location, which is available at a 0.5°x0.5° latitude-longitude spatial resolution.

For our historical baseline, we used data from 2015–2019, a five-year period encompassing the Zika outbreak in Brazil. For our forecast, we used 30-year projections, i.e., projected temperature data for the years 2045–2049. For the temperature projections, we use four SSP (Shared Socioeconomic Pathways) climate scenarios: SSP126, SSP245, SSP370, and SSP585 [30]. These scenarios represent different climate-relevant levels of socioeconomic development (taking into consideration factors like sustainable consumption, protection of vulnerable land, fertility and rates) and their corresponding greenhouse gas concentrations. Here, increasing climate change severity corresponds to increasing numbers, where SSP585 corresponds to fossil-fueled developed while the SSP126 would require substantial mitigation efforts on a global level to achieve. The GFDL-ESM4 model provides temperatures for previous years for each of the four SSP scenarios, starting in 2015, (i.e., each scenario has different historical temperature data for those years). We use the SSP585 scenario for our historical temperatures (most closely corresponding to RCP (Representative Concentration Pathway) 8.5 from CMIP-5), because this trajectory is thought to most closely align with the carbon dioxide emissions from those years [31, 32].

We summarized the historical and projected temperature data in two ways using period cubic B-splines. First, we obtain both a baseline year-round temperature dataset by averaging the daily temperatures from our 5-year dataset (2015–2019), and a 2045–2049 projected year-round temperature dataset from the averages of the years 2045–2049 (see S1 Appendix). By smoothing over the 5 years, we projected average climate and smooth any anomalies that occur in the projections for 2045–2049, giving an estimation of overall changes in risk by the second half of the 2040s as compared to the recent past. Second, to better capture the year-to-year variation, we also fit the periodic splines to each of the five years separately for both the historical and projected temperatures to better understand reasonable likely deviation from the mean projection (see S1 Appendix). The period cubic B-splines were fit to the temperature data using the pbs package [33] in R (v4.0; R Foundation for Statistical Computing; Vienna, Austria).

### Infectious disease transmission model

We modified an existing vectorborne infectious disease transmission model [21] to include birth and death processes. The basic model structure comprises an SLIR (susceptible, latent, infectious, recovered) model for human transmission and SLI model for mosquito transmission, using standard exponential birth and death processes for the human population. The model tracks the numbers of susceptible *S*_*h*_, latently infected *L*_*h*_, infectious *I*_*h*_, and recovered *R*_*h*_ humans (with total human population *N*_*h*_), as well as the number of susceptible *S*_*m*_, latently infected *L*_*m*_, and infectious *I*_*m*_ mosquitoes. Our model includes three temperature-independent, human parameters: the birth/death rate *μ*_*h*_, the transition rate from latency to infectiousness *σ*_*h*_ (which we assumed to be two days less than the intrinsic incubation period, as infectiousness precedes symptom onset [34]), and the recovery rate *γ*. The birth and death rates *μ*_*h*_ were fixed to single value based on current life expectancy for this analysis rather than projected; because the model is focused on epidemic potential (see below) rather than simulation, the results are not sensitive to these values (impacting the results only in the probability that that an latent or infected individual may die before recovery).

Our model also includes eight temperature-dependent (*T*) *A. aegypti* mosquito parameters, five of which are independent of the pathogen: biting rate *a*(*T*), the number of eggs laid per day *ϵ*(*T*), the probability of egg to adult survival *θ*(*T*), the egg to adult development rate *ρ*(*T*), and the adult mosquito mortality rate *μ*_*m*_(*T*). One temperature dependent mosquito parameter not included in our eight parameters is the carrying capacity *K*(*T*), which is the maximum number of mosquitoes that the environment can sustain. This parameter can be modeled as function of the other vector parameters [21] (see S1 Appendix) and therefore does not appear in the *ℛ*_0_(*T*) formula we derive.

The three additional temperature-dependent parameters depend on the specific pathogen: the extrinsic incubation rate, that is the latency to infectiousness rate *σ*_*m*_(*T*), the per bite probability of pathogen transmission from mosquito to human *π*_*mh*_(*T*), and the per bite probability of pathogen transmission from human to mosquito *π*_*hm*_(*T*). We define vector competence as the product of *π*_*mh*_(*T*) and *π*_*hm*_(*T*), denoted (*π*_*hm*_*π*_*mh*_)(*T*). For Zika we have temperature dependent estimates for the vector competence product but not the constituent parameters.

Like Mordecai et al [17, 35], the temperature dependence of these 8 parameters are described by one of four formulas: a Brière 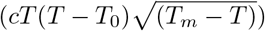, quadratic (*c*(*T − T*_*m*_)(*T − T*_0_)), inverse quadratic (*c*(*T − T*_*m*_)(*T − T*_0_))^*−*1^, or constant *c*, as appropriate for the shape of the relationship in the data (Table 1). *T*_0_ and *T*_*m*_ are the minimum and maximum temperatures for which a given parameter takes on a non-zero value. The parameter *c* is fit to the data [35, 36]. Plots of the temperature dependence of the biting rate *a*, the extrinsic incubation rate *σ*_*m*_, and the vector competence (*π*_*hm*_*π*_*mh*_) are given in the Figure S3, distinguishing between dengue and Zika where appropriate.

**Table 1.** Parameters of the temperature-dependent vectorborne arbovirus disease transmission model. *†*: see supplementary material.

| Temperature-dependent parameters (Zika and dengue) |  |  |  |  |  |  |
| --- | --- | --- | --- | --- | --- | --- |
| Parameter | Definition | Source | Function | $T_0$ | $T_m$ | $c$ |
| $a$ | biting rate ($\text{day}^{-1}$) | [35] | Brière | 13.35 | 40.08 | 2.02E-4 |
| $\epsilon$ | eggs laid per female ($\text{day}^{-1}$) | [35] | Brière | 14.58 | 34.61 | 8.56E-3 |
| $\theta$ | probability of mosquito egg to adult survival | [35] | Quadratic | 13.56 | 38.29 | -5.99E-3 |
| $\rho$ | mosquito egg to adult development rate ($\text{day}^{-1}$) | [35] | Brière | 11.36 | 39.17 | 7.86E-5 |
| $\mu_m$ | adult mosquito mortality rate | [35, 36] | Inverse | 8.53 | 38.07 | -1.68E-1 |
| Temperature-dependent parameters (Zika) |  |  |  |  |  |  |
| $\sigma_m^z$ | virus extrinsic incubation rate | [36] | Brière | 18.27 | 42.31 | 1.74E-4 |
| $(\pi_{hm}\pi_{mh})^z$ | vector competence | [36] | Quadratic | 22.72 | 38.38 | -3.54E-3 |
| Temperature-dependent parameters (dengue) |  |  |  |  |  |  |
| $\sigma_m^d$ | virus extrinsic incubation rate | [35]† | Brière | 10.68 | 43.09 | 6.91E-5 |
| $\pi_{mh}^d$ | probability of transmission to human (per bite) | [35] | Brière | 17.05 | 35.83 | 8.49E-4 |
| $\pi_{hm}^d$ | probability of transmission to vector (per bite) | [35] | Brière | 12.22 | 37.46 | 4.91E-4 |
| Temperature-independent parameters (Zika and dengue) |  |  |  |  |  |  |
| $\mu_h$ | human birth/death rate ($\text{day}^{-1}$) | [37] | Constant | — | — | $(75.7 \times 365)^{-1}$ |
| $\frac{N_m}{N_h}$ | Ratio of *A. aegypti* to humans | [38] | Constant | — | — | 9.75E-1 |
| Temperature-independent parameters (Zika) |  |  |  |  |  |  |
| $\sigma_h^z$ | human latency rate ($\text{day}^{-1}$) | [34, 39] | Constant | — | — | 1/4 |
| $\gamma^z$ | human recovery rate ($\text{day}^{-1}$) | [34, 39] | Constant | — | — | 1/5 |
| Temperature-independent parameters (dengue) |  |  |  |  |  |  |
| $\sigma_h^d$ | human latency rate ($\text{day}^{-1}$) | [34] | Constant | — | — | 1/4 |
| $\gamma^d$ | human recovery rate ($\text{day}^{-1}$) | [34] | Constant | — | — | 1/5 |

Here we use different values of lifespan and extrinsic incubation estimates from Mordecai et al [35]. Because the mosquito mortality rate should largely be independent of the pathogen, we merge the data from [35] and [15] to generate a compiled temperature-dependent mosquito mortality *μ*_*m*_(*T*). Maximum likelihood estimates for the parameters *c, T*_0_, and *T*_*m*_ were obtained assuming mosquito lifetimes were Poisson distributed (see supplementary material). Similarly, the extrinsic incubation rate was refit to exclude sources from papers which studied other arboviruses such as Yellow Fever. We parameterize the number of mosquitoes (*N*_*m*_) and the number of humans (*N*_*h*_) as a single parameter, 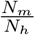, corresponding to the density of mosquitoes (i.e., the number of mosquitoes per human).

The parameters are summarized in Table 1, and the model equations are given below.

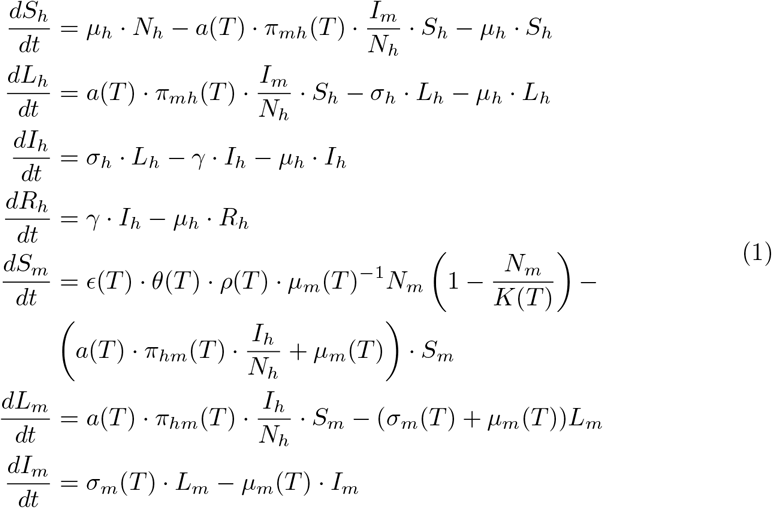

### Basic reproduction number

The basic reproduction number *ℛ*_0_ is a measure of the epidemic potential of an infectious disease system [40, 41]. It represents the expected number of secondary infections caused by a single infectious case over their infectious period in an otherwise susceptible population. If *ℛ*_0_ *>* 1, an epidemic is expected to grow and if *ℛ*_0_ *<* 1, an epidemic is expected to die out. *ℛ*_0_ is an appropriate metric for our our projections because there is too much uncertainty what specific circulation patterns will be over time and in population-level immunity to project specific outbreak dynamics in 30 years. Our approach instead focuses on transmission potential. Even if there is substantial population immunity suppressing Zika and dengue circulation, understanding transmission potential is still useful and can inform arboviral disease potential more broadly.

In the context of vectorborne disease systems, there is some subtlety to the interpretation of *ℛ*_0_: strictly speaking, a disease generation-based *ℛ*_0_, as derived by the next generation matrix (NGM) [42, 43] and denoted below as 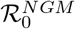, treats hosts (humans) and vectors (mosquitoes) as equally important, essentially taking the mean of human-to-vector infections and vector-to-human infections. Because we observe human cases, only, it is usually preferable and more interpretable to use the expected number of new human infections per infectious human, namely 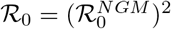. This formulation is consistent with classic approaches [44]. We use the next generation method to derive a formula for 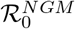 and thus this latter temperature-dependent *ℛ*_0_(*T*) for our model (see S1 Appendix).

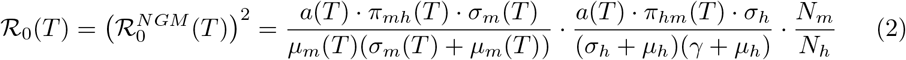

Incorporating the virus-specific parameter values into this expression, we derive values for Zika,

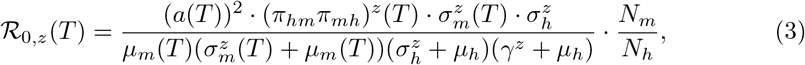

and dengue,

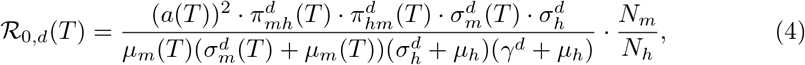

as a function of the pathogen-specific, temperature dependent parameters.

## Results

### Temperature-dependent basic reproduction numbers

The temperature-dependent shape of the *ℛ*_0_(*T*) curve is similar for Zika and dengue (Figure 2). For example, the peak *ℛ*_0_ occurs at approximately 30.5°*C*. One the other hand, *ℛ*_0_ increases above 1 at a cooler temperature for dengue compared to Zika (23°*C* vs 25°*C* respectively), and the peak *ℛ*_0_ value is greater for dengue than Zika (6.8 vs 2.7). The greater *ℛ*_0_ for dengue is primarily driven by vector competence (the probability of transmission to human times the probability of transmission to vector, see supplementary material). To a lesser degree, the shorter extrinsic incubation period of DENV (Dengue virus) also contributes to its larger *ℛ*_0_. Note that the *ℛ*_0_ metric considers a fully susceptible population, and the effective reproduction numbers for real populations decrease proportionally to the fraction of the population that is immune.

**Fig 1.**
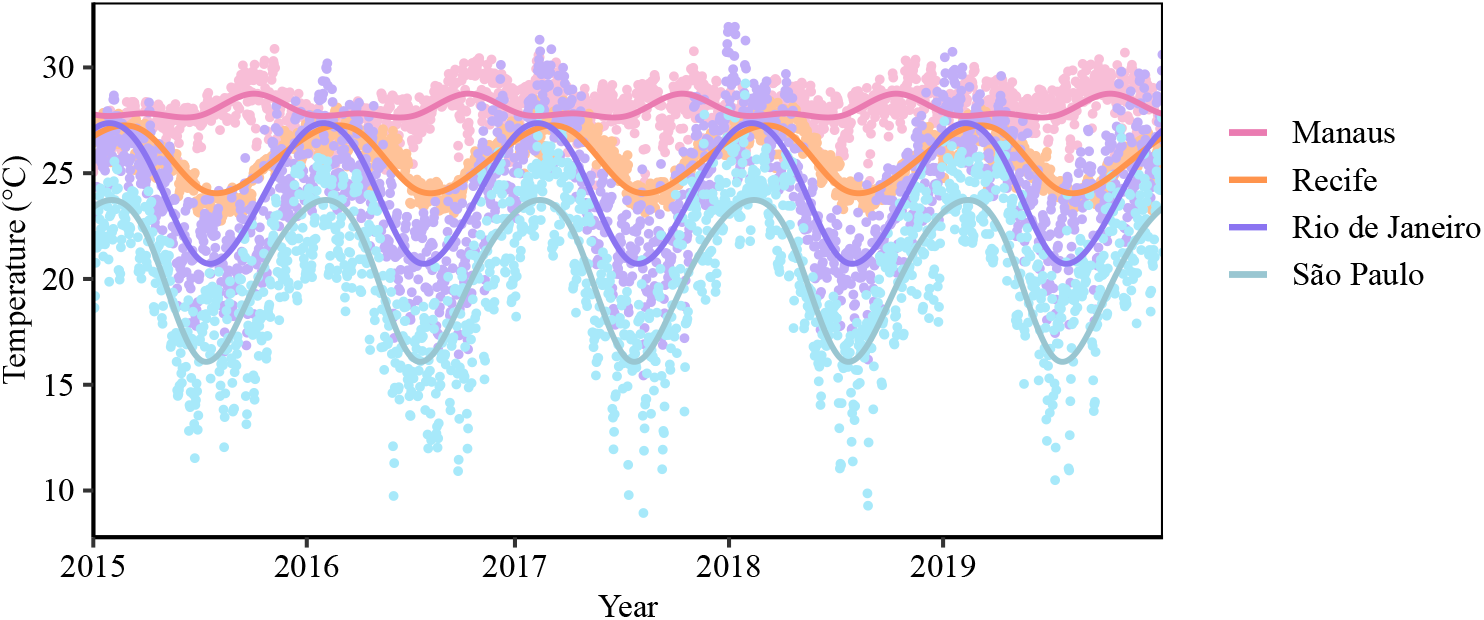
Daily temperature in 4 Brazilian cities, 2015–2019. Periodic cubic spline models are fit to the data for Manaus, Recife, Rio de Janeiro, and São Paulo to develop mean seasonal temperature models.

**Fig 2.**
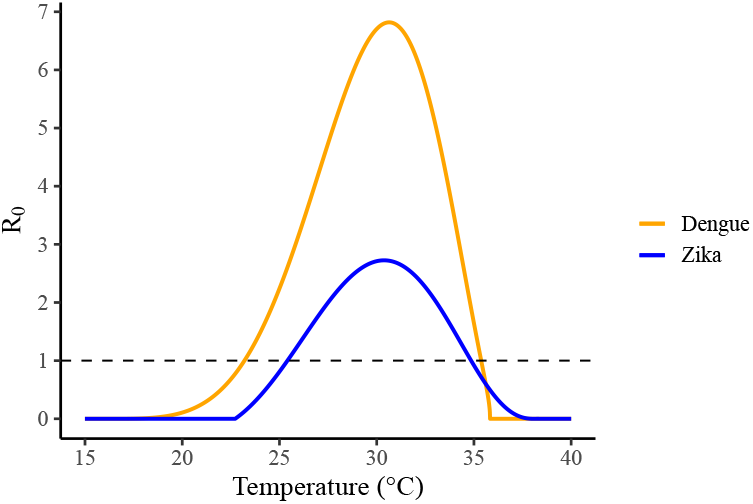
Temperature-dependent *ℛ*_0_(*T*) for Zika and dengue.

### Risk projections based on 5-year temperature data

The climate change scenarios project a year-round increase of *ℛ*_0_ by 2045–2049, with varying degrees of difference among the risk projections between the specific SSP scenarios (Figure 3). Exceptions include the extreme temperatures seen in the warm season in Manaus and the cool season in Rio de Janeiro and São Paulo. For Manaus, we predict the annual Zika *ℛ*_0_ range, currently 2.1–2.5, to shift to 2.3–2.7, for Recife we project the range to shift from 0.4–1.9 to 0.6–2.3, for Rio de Janeiro to shift from 0–1.9 to 0–2.3, and for São Paulo to shift from 0–0.3 to 0–0.7. The increase in *ℛ*_0_ is not uniform throughout the year as can be seen in the graphs for Rio de Janeiro in particular, where the *ℛ*_0_ value increases by a far larger amount during the months of October through April than it does earlier in the year (*ℛ*_0_ remains 0 throughout the summer months in all scenarios, but increases as high as 0.8 in the fall and winter months). To a lesser extent, *ℛ*_0_ increases are also non-uniform for Recife (increasing around 0.1 earlier in the year and as high as 0.5 by late Summer). These effects are due to a combination of the non-uniform temperature changes in the temperature projection data over the year and the non-linearity of the *ℛ*_0_ formula. We see some minor attenuation of risk because of higher temperatures across the risk projections in the warmest months in Manaus, where temperatures are projected to reach just above 35°*C* in the SSP585 scenario. However, in this scenario, the peak risk still far surpasses that of the baseline risk, occurring at two different times in the year corresponding to the bookends of the observed dip in risk (around September and November).

**Fig 3.**
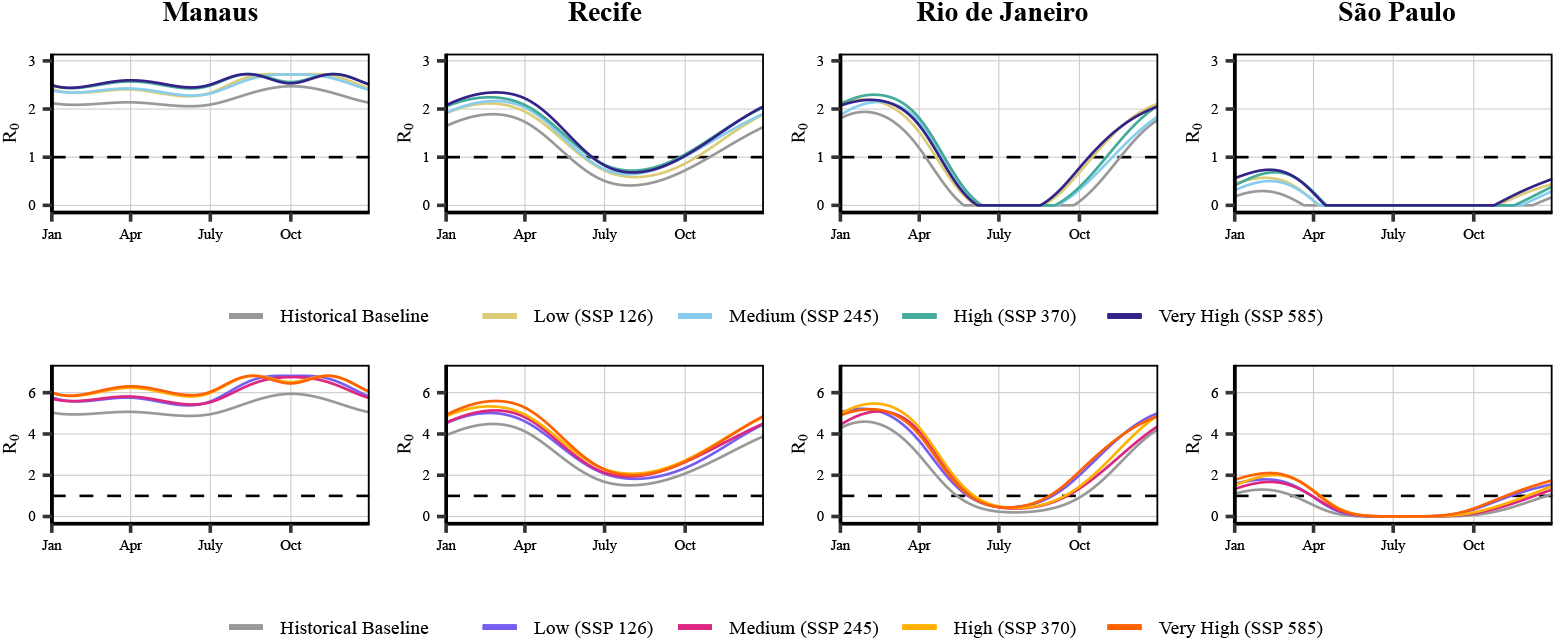
Projection of seasonal epidemic potential by 2045-2049 *ℛ*_0_(*T* (*t*)). Projections are given for Zika (top row) and dengue (bottom row) for each city and climate change scenario.

Our baseline risk estimates for Rio de Janeiro and Recife suggest that the current risk season for Zika, i.e., the time for which *ℛ*_0_ *>* 1, is late fall through spring which is largely consistent with data from the 2015–16 outbreak [45, 46]. Dengue follows a similar trend, but with a longer risk season. Our risk projections suggest that the arbovirus risk season for Rio de Janeiro will increase by approximately 2–3 months by 2045–2049 and that the Zika risk seasons in Recife will increase by around 2 months. In São Paulo, the R0 for dengue more reliably sits above 1 during the beginning of the year in our projections for 2045–2049, peaking at 2.1 in SSP585, nearly double the peak R0 value from the baseline.

### Risk projections based on individual-year data

Our risk projections based on individual year data (Figure 4) highlight the heterogeneity of the Zika risk projections from year-to-year (the corresponding figure for dengue can be found in S1 Appendix). The projections for Manaus contrast with the projections from the other three cities, which still show largely consistent increase in disease risk throughout the year for each year. For example, Manaus sees a sharp decrease in risk in fall 2045 for SSP585, demonstrating potentially erratic shifts in peak risk seasons for this city to earlier in the year. As it is likely that outbreaks in Recife and Rio de Janeiro fuel epidemic potential in Manaus and vice versa (i.e., through travel between the cities), this closer alignment of peak risk season between the cities may cause a problematic interaction.

**Fig 4.**
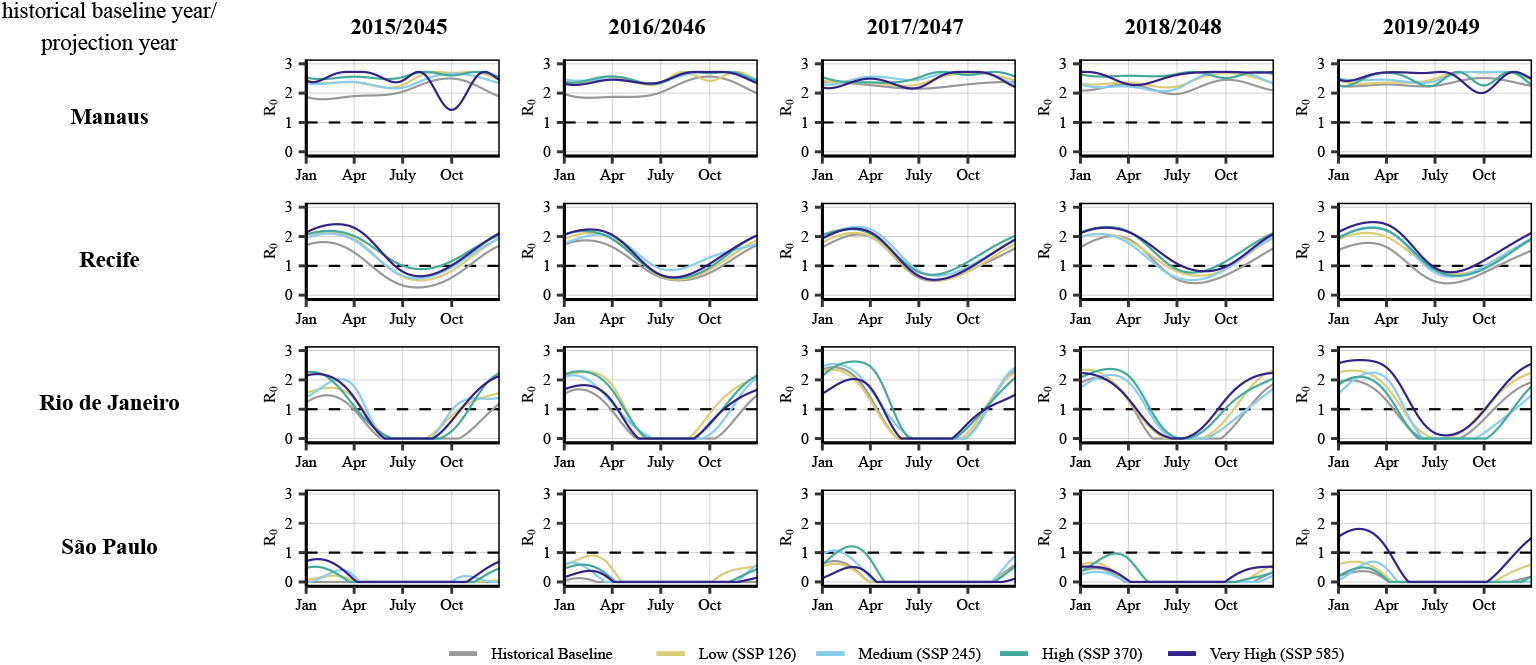
Year-to-year variation of seasonal epidemic potential *ℛ*_0_(*T* (*t*)) projections for Zika. Projections of the seasonal temperature in each of 2015–2019 under each climate change scenario demonstrate year-to-year heterogeneity in projected risk.

São Paulo’s risk is also highly variable between the different years and SSP scenarios, highlighting important distinctions between each of the scenarios. For example, our projections show a dramatic difference in the year 2049 for the SSP585 scenario compared to the other scenarios. These results suggest that the year-to-year heterogeneity in temperature and thus on arbovirus disease risk in the future will likely depend on regional climate factors and year-specific weather patterns.

## Discussion

Zika and dengue’s temperature profile for *ℛ*_0_ peaks at a relatively high temperature, around 30°*C*; therefore, prior work has suggested that climate change will both increase their transmission potential and geographic extent of transmission [19]. When examining the impact of climate change projections on Brazil, we find general agreement with this conclusion but also find variability across different climatic regions within the country. This variability across different climate zones is evident in Figure 4, which shows that Manaus is a region on the cusp of experiencing a decrease in arbovirus risk at certain times of the year in certain years, while both Recife and Rio de Janeiro show large increases in risk throughout the year. In places like Recife and Rio de Janeiro, we project the extension of the risk season. In São Paulo, a city that lies on borderline of reliable *A. aegypti* suitability [3, 47, 48] we see that it is likely that future arboviral risk will depend on how specifically the climate changes. These results highlight that transmission is likely to expand into geographic regions with climates that currently have only borderline conditions for transmission. Regions with current temperatures that are too cold to sustain year-round transmission will become increasingly vulnerable to newly seeded outbreaks sparking seasonal epidemics.

Temperature-dependent *ℛ*_0_(*T*) curves, used here and in other studies, indicate that there is a potential for increasing temperatures to have a protective effect. The curves for dengue and Zika begin to decrease sharply after they peak at around 30°C, both decreasing to 1 by around 35°C (95°F). Of the four cities in the analysis, Manaus is the only city to reach this peak temperature, and even in the high emission scenario, the maximum temperature in our projections is only briefly above 30°*C* at the beginning of October. Thus, even in regions with warmer tropical rain forest climates like Manaus, our results show that in most regions climate change is not likely to have a substantial or consistent protective effect on arbovirus transmission.

Over the past decades, numerous studies have looked at the impact of temperature changes on vector-borne disease transmission [3, 5, 13, 16, 20, 49–51]. There is general consensus among these studies that both dengue and Zika will spread into areas that are becoming increasingly suitable for transmission (e.g., the Southeastern United States) and that risk will increase in currently endemic areas. The temperature-dependence of mosquito-borne disease transmission is a complicated mix of multiple processes, each of which increases, then decreases with increasing temperature. Several studies concur that 26–29°C is the optimal temperature window for arbovirus transmission [35, 36, 52, 53]. Zika and dengue lie on the higher end of this range [20], at around 30°*C*, which is consistent with our estimates. Our *ℛ*_0_(*T*) estimates also span ranges that are consistent with empirical estimates: a systematic literature review on the basic reproduction numbers for dengue and Zika found the *ℛ*_0_ of Zika (mean 3.0) to be lower than the *ℛ*_0_ of dengue (mean 4.3) in tropical climates, and our estimated values are well within the substantial variation in individual study estimates [54]. Our *ℛ*_0_ for Zika is just below 3.0 even at its highest value.

The variability between the various SSP scenarios seen in Figure 4 along with steepness of the temperature-dependent *ℛ*_0_ curves (Figure 2) underscore the severe consequences of small deviations in temperature projections with regards to arbovirus risk. That is, our ability to control our emissions to prevent even small temperature increases could have massive benefits relating to mosquito-borne illnesses. That being said, by 2045–2049, even the best-case scenario (SSP126) corresponds to both lengthening of the risk season—particularly for Rio de Janeiro—and increases in overall disease pressure, indicating that international climate protection policy must be accompanied with national-level preparedness including increased surveillance, and diagnostic and treatment capacity.

For this analysis, we chose to focus on epidemic potential through the basic reproduction number. Previous work has demonstrated that beyond epidemic potential, there will be likely differences in epidemic dynamics, such as epidemic length, peak size, and final size [21]. However, for the long-term projections we provide here, *ℛ*_0_ is appropriate—–there are too many unknowns in terms of what population-level immunity will look like to make reasonable projections of specific dynamics. Indeed, it is unclear whether these arboviruses will be circulating in the coming decades and whether new pathogens will emerge. Thus, one strength of this study is the side-by-side comparison of dengue and Zika risk, which gives a broader look at arbovirus epidemic potential, regardless of the specific pathogen. Another strength is the applicability of our results to other areas; the temperature-dependent *ℛ*_0_(*T*) we derived here could be implemented for other areas of interest. Finally, our study also uses a fine temporal granularity, which gives us the ability to provide a more in depth understanding of year-round dynamics and investigate arbovirus risk as a dynamic value that changes over the course of a month or year.

However, our work is limited by its sole focus on temperature. Climate change is likely to impact humidity and rainfall, and population density will also likely change in the future. We do not account for these factors in our projections. Due to its extreme variability, precipitation is generally thought to be a more complicated climactic factor to project than temperature [55, 56] and including this would have introduced tremendous uncertainty to the results, even using state-of-the-art model projections. Another limitation is the uncertainty in population birth rates and changes in the mosquito-to-human population density ratio. Changes in these parameters may impact the epidemic potential of Zika and other arboviruses.

## Conclusion

Our work provides a useful baseline for future planning to mitigate health impacts due to climate change. It contributes to the larger literature of climate change health impacts by exploring the likely heterogeneities in these health impacts both across climatic regions within a country and from year-to-year. International cooperation will be needed to mitigate emissions and their impacts up to and beyond 2045–2049. Specifically, we should be developing our public health preparedness to offset increases in transmission pressure due to climate. Transmission models coupled with climate forecasts can provide the needed guidance on how best to develop a preparedness infrastructure that will be resilient to climate change. Greater flexibility and adaptability of arbovirus response and prevention may be necessary to accommodate spatial and temporal heterogeneity in risk projections, especially in a country with as much climatic diversity as Brazil.

## Data Availability

All data were derived from publicly available sources.

https://doi.org/10.48364/ISIMIP.581124.1

## S1 Appendix. Supporting information

In the supporting information, we provide the periodic spline fits to the individual years in each city, provide the periodic spline fits to the 5-year temperatures under each of the climate change scenarios in each city, discuss the temperature-dependent mosquito carrying capacity, give the temperature-dependent parameter models as well as the fits to the data were applicable, and provide the individual-year risk projections for dengue (analogous to Figure 4).

